# Large scale phenotyping of long COVID inflammation reveals mechanistic subtypes of disease

**DOI:** 10.1101/2023.06.07.23291077

**Authors:** Felicity Liew, Claudia Efstathiou, Sara Fontanella, Matthew Richardson, Ruth Saunders, Dawid Swieboda, Jasmin K. Sidhu, Stephanie Ascough, Shona C. Moore, Noura Mohamed, Jose Nunag, Clara King, Olivia C. Leavy, Omer Elneima, Hamish J.C. McAuley, Aarti Shikotra, Amisha Singapuri, Marco Sereno, Victoria C Harris, Linzy Houchen-Wolloff, Neil J Greening, Nazir I Lone, Matthew Thorpe, A. A. Roger Thompson, Sarah L. Rowland-Jones, Annemarie B. Docherty, James D. Chalmers, Ling-Pei Ho, Alexander Horsley, Betty Raman, Krisnah Poinasamy, Michael Marks, Onn Min Kon, Luke Howard, Daniel G. Wootton, Jennifer K. Quint, Thushan I. de Silva, Antonia Ho, Christopher Chiu, Ewen M Harrison, William Greenhalf, J. Kenneth Baillie, Malcolm G. Semple, Rachael A. Evans, Louise V. Wain, Christopher Brightling, Lance Turtle, Ryan S. Thwaites, Peter J.M. Openshaw, ISARIC4C Investigators and the PHOSP-COVID collaborative group

## Abstract

One in ten SARS-CoV-2 infections result in prolonged symptoms termed ‘long COVID’, yet disease phenotypes and mechanisms are poorly understood. We studied the blood proteome of 719 adults, grouped by long COVID symptoms. Elevated markers of monocytic inflammation and complement activation were associated with increased likelihood of all symptoms. Elevated IL1R2, MATN2 and COLEC12 associated with cardiorespiratory symptoms, fatigue, and anxiety/depression, while elevated MATN2 and DPP10 associated with gastrointestinal (GI) symptoms, and elevated C1QA was associated with cognitive impairment (the proteome of those with cognitive impairment and GI symptoms being most distinct). Markers of neuroinflammation distinguished cognitive impairment whilst elevated SCG3, indicative of brain-gut axis disturbance, distinguished those with GI symptoms. Women had a higher incidence of long COVID and higher inflammatory markers. Symptoms did not associate with respiratory inflammation or persistent virus in sputum. Thus, persistent inflammation is evident in long COVID, distinct profiles being associated with specific symptoms.

## Main

One in ten SARS-CoV-2 infections results in long COVID or Post-acute sequelae of COVID-19 (PASC) resulting in an estimated 17 million Europeans experiencing prolonged symptoms during the first two years of the pandemic.^1^ SARS-CoV-2 continues to circulate and long COVID remains common, even after mild infection with more recent variants.^2, 3^ It is likely long COVID will continue to be a substantial cause of long-term ill health, requiring targeted management based on specific disease phenotypes and underlying mechanisms.

There are reports of persistent inflammation in adults with long COVID,^4–7^ but studies have been limited by size, timing of samples or number and breadth of immune mediators measured, leading to inconsistent associations with symptoms. The PHOSP-COVID study recently reported the plasma proteome of 626 adults, showing inflammation in those with very severe long COVID symptoms based on clustering, utilising measures of breathlessness, fatigue, mental health, cognitive impairment, and physical performance.^8, 9^ However, it is unclear if there are changes in the proteome specific to symptom type compared to recovered controls. It is therefore unknown if there are common pathways of inflammation driving long COVID symptoms or if different patterns of inflammation underly specific clinical subtypes, requiring tailored therapeutic approaches. Understanding the immunological mechanisms that underlie long COVID has been highlighted as a top research priority by patients, clinicians and scientists.^10^

Many long COVID symptoms have been described, most commonly breathlessness, fatigue, memory impairment and gastrointestinal (GI) disturbance.^11, 12^ There are reports of autoantibodies in those with respiratory or GI symptoms, whilst Epstein-Barr Virus reactivation has been associated with fatigue and neurological symptoms,^5, 13–15^ suggesting that distinct mechanisms might cause different symptom types and that these might be revealed by analysis of inflammatory markers.

In this prospective multicentre study, we measured 360 plasma proteins in 719 adults six months after hospitalisation for COVID-19, confirmed either clinically or by PCR (n=621). Within our cohort, including the 626 patients previously analysed,^9^ 250/719 (35%) reported full recovery (“Recovered”), the remaining 469 (65%) reported persistent symptoms consistent with long Covid (fig 1A; table 1). Using a penalised logistic regression model (PLR) to explore the influence of patient demographics and immune mediators on symptoms, we found female sex to predict all symptoms and to be the strongest predictor of GI (Odds Ratio [OR]=1.13) and cardiorespiratory symptoms (OR=1.17; fig 1B–F). Confidence intervals are not appropriately derived from PLR analysis and are therefore not reported, however repeated cross-validation was used to estimate uncertainty associated with PLR outputs (methods and supplemental). Pre-existing conditions that might predispose to adverse outcomes (e.g., chronic lung disease and prior cardiorespiratory symptoms) were a risk factor for all symptoms (except GI); age and acute disease severity were not associated with any symptom. We did not include ethnicity as a covariate because it is not an independent risk factor in this cohort.^11^

**Figure 1.**
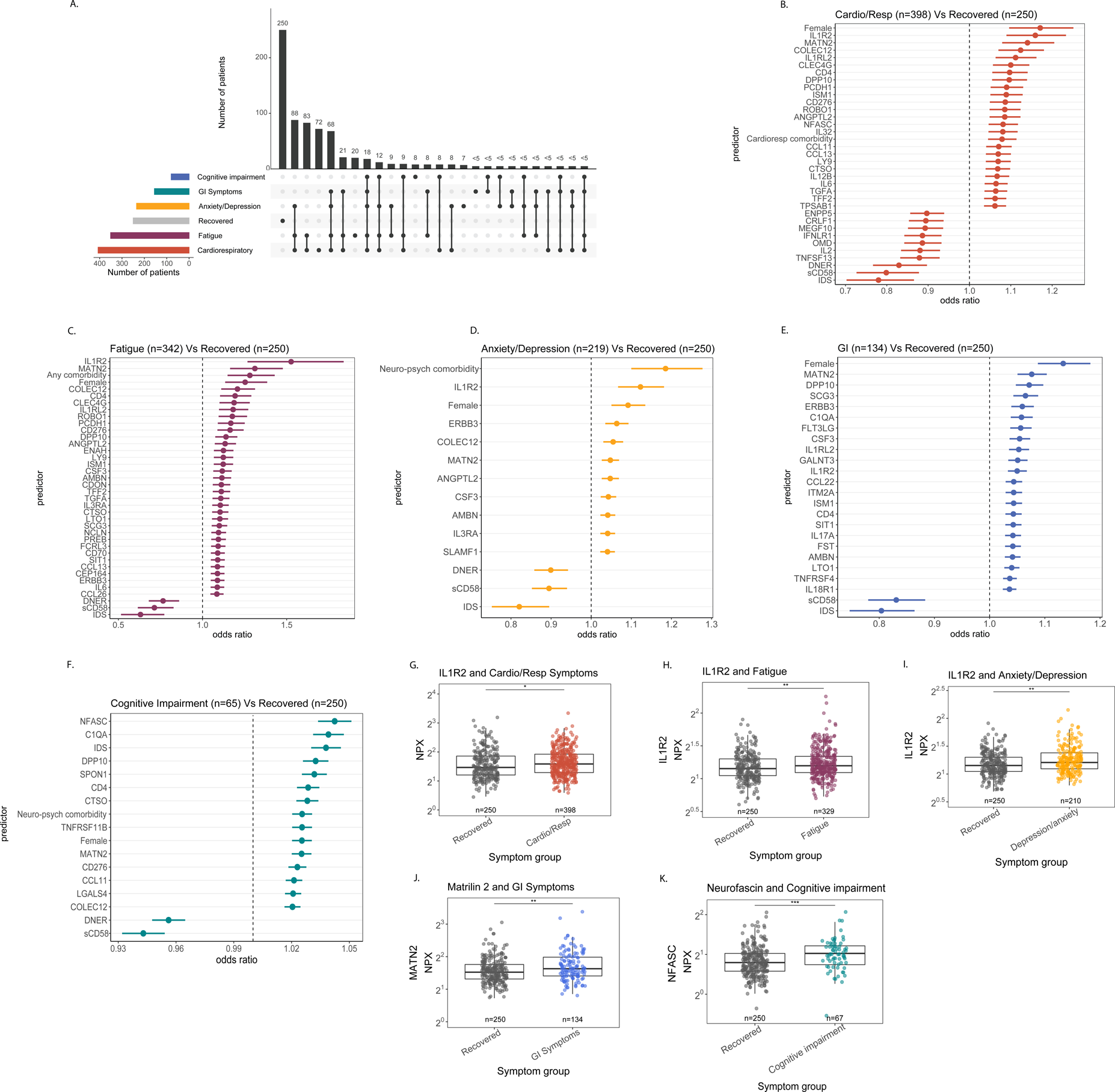
Prevalence of long COVID symptoms and their associated protein signature. 719 patients were recruited 6 months after COVID-19 hospitalisation and divided according to their long COVID symptom group (A). The horizontal coloured bars represents the number of patients in each symptom group. The vertical black bars represent the number of patients in each symptom combination group. To prevent patient identification, where less than 5 patients belong to a combination group, this has been represented as ‘<5’. Within the cohort 250 patients reported feeling recovered and were used as controls (grey horizontal bar). Forest plots of protein signatures associated with cardiorespiratory symptoms (B), fatigue (C), anxiety/depression (D), GI symptoms (E) and cognitive impairment (F). Odds ratios were derived from PLR coefficients. Sex, age, acute disease severity and comorbidities were included as covariates. Confidence intervals cannot be derived from PLR and error bars represent the median accuracy of the model. The distribution of inflammatory proteins associated with the highest odds of each outcome and compared to protein levels in the recovered group are shown (G–K). Median values were compared between groups using the Wilcoxon signed-rank test. * = p<0·05, ** = p<0·01, *** = p<0·001, ****= p<0·0001.

**Table 1.** Cohort demographics. The demographics of each symptom group and recovered controls are shown. Differences between groups were compared using chi-squared or ANOVA as appropriate. Data are n (%) or mean (SD). C-Reactive Protein (CRP) levels represent those measured contemporaneously with inflammatory protein measurements reported in this study.

|  |  | **GI** | **Fatigue** | **Cardio-respiratory** | **Anxiety/depression** | **Cognitive impairment** | **Recovered** | ***p*** |
| --- | --- | --- | --- | --- | --- | --- | --- | --- |
| **Age at admission** | Years (SD) | 57.72 (11.48) | 56.57 (11.07) | 57.08 (11.37) | 56.36 (10.84) | 59.24 (12.82) | 58.92 (13.72) | *p*=0.046<br>* |
| **Sex** | Female<br>N (%) | 68 (53%) | 143 (47%) | 161 (43%) | 89 (45%) | 24 (42%) | 55 (27%) | *p*<0.001<br>**** |
| **Ethnicity** | White | 110 | 300 | 331 | 193 | 50 | 197 | *p*=0.09 |
|  | South Asian | 14 | 26 | 38 | 16 | 7 | 46 |  |
|  | Black | 8 | 16 | 25 | 11 | 7 | 10 |  |
|  | Mixed/Other | 8 | 24 | 22 | 16 | 7 | 17 |  |
| **WHO Clinical Progression Scale** | Class 3-4 | 41 | 83 | 88 | 45 | 18 | 45 | *p*=0.28 |
|  | Class 5 | 45 | 107 | 124 | 74 | 21 | 115 |  |
|  | Class 6 | 27 | 78 | 89 | 55 | 11 | 57 |  |
|  | Class 7-9 | 27 | 98 | 115 | 62 | 21 | 50 |  |
| **CRP** | Mean (SD) | 5.24 (2.62) | 5.36 (2.30) | 5.10 (2.24) | 5.69 (2.47) | 4.52 (2.17) | 4.63 (1.67) | *p*=0.71 |
| **Comorbidities** | Mean (SD) | 2.9 (2.62) | 2.675 (2.3) | 2.553 (2.24) | 2.911 (2.47) | 2.493 (2.17) | 1.554 (1.67) | *p*=<0.0001<br>**** |

To study the association of peripheral inflammation with symptoms, 360 immune mediators were measured from plasma and included as covariates in the model. PLR enabled adjusted coefficients, accounting for the number of covariates included and collinearity between variables, as appropriate for high dimensional data.^16^ Given the overlap between symptom groups (fig 1A) and allowing for collinearity, PLR enabled specific predictive mediators for each symptom group to be distinguished.

Mediators suggestive of persistent monocytic inflammation were associated with all symptoms (fig 1B–F). Specifically, elevated IL1R2 and/or Matrilin-2 (MATN2) were consistently associated with the highest odds of all symptoms except cognitive impairment (cardiorespiratory IL1R2 OR=1.16; fatigue IL1R2 OR= 1.53; anxiety/depression IL1R2 OR=1.13; GI MATN2 OR=1.08). IL1R2 is expressed by monocytes and macrophages and modulates IL1-driven inflammation.^17^ MATN2 is an extracellular matrix (ECM) protein which promotes inflammation by activating toll-like receptors and enhancing monocyte infiltration into tissues.^18, 19^ Although the effect was smaller, CSF3 (G-CSF, which promotes inflammation by stimulating neutrophil production), was associated with GI symptoms (OR=1.05), fatigue (OR=1.12) and anxiety/depression (OR=1.05) (fig 1C–E).^20^ IL-6 was also associated with increased odds of cardiorespiratory symptoms (OR=1.06) and fatigue (OR=1.09) (fig 1B&C). These findings are consistent with previous studies suggesting IL1- and myeloid-driven inflammation contribute towards the pathogenesis of depression,^21, 22^ as well as two previous studies finding IL1-driven and monocytic inflammation in adults with long COVID.^23, 24^

Although these findings are indicative of systemic inflammation, C-reactive protein levels, a commonly used clinical marker of inflammation, were not significantly different between long COVID and recovered groups (table 1). Elevated sCD58 (lymphocyte-function antigen 3, an immunosuppressive factor),^25^ was associated with reduced odds of all long COVID symptoms and this was most pronounced for fatigue (OR=0.71) and cardiorespiratory symptoms (OR=0.79) (fig 1B–F). Collectively, our findings suggest systemic monocytic inflammation underlies long COVID, and that resolution of this may be associated with recovery.

Collectin-12 (COLEC12) was associated with increased odds of experiencing cardiorespiratory symptoms (OR=1.12), anxiety/depression (OR=1.06) and fatigue (OR=1.21) (fig 1B–D). COLEC12 can initiate inflammation in tissues by activating the alternative complement pathway and by modulating leucocyte recruitment.^26, 27^ Whilst COLEC12 was not associated with GI symptoms and only weakly associated with cognitive impairment (OR=1.02), C1QA did predict these symptoms (fig 1E&F). C1QA is a component of the complement system, initiating inflammation via the classical pathway.^28^ Notably, C1QA was associated with the third highest odds of cognitive impairment (OR=1.04), and has been implicated in the pathogenesis of chronic neuroinflammation in Alzheimer’s disease.^29^ Although differences were observed between symptom groups, monocytic inflammation and complement activation were common to all adults with long COVID.^29, 30^

Multiple mechanisms for long COVID have been suggested including autoimmunity, thrombosis, SARS-COV-2 persistence or latent virus reactivation.^5, 24, 31^ The protein signatures we observed would be consistent with these mechanisms since both infection and autoantibody production can activate complement.^28, 32^ Unregulated complement activation drives systemic inflammation, thrombosis and tissue damage providing a plausible explanation for some long COVID symptoms and a potential target for therapeutic trials.^33^

Whilst several common proteins predicted cardiorespiratory symptoms, anxiety/depression and fatigue (the largest combination group; n=88), the protein signature associated with GI symptoms and cognitive impairment appeared distinct (fig 1E&F). MATN2, Dipeptidyl peptidase 10 (DDP10) and Secretogranin 3 (SCG3) were associated with the highest odds of GI symptoms (MATN2 OR=1.08; DPP10 OR=1.07; SCG3 OR=1.06). DDP10 is a membrane protein which can modulate tissue inflammation, and increased expression of *DPP10* is associated with Ulcerative Colitis, suggesting that post-COVID GI symptoms may result from enteric inflammation.^34, 35^ Additionally, elevated SCG3 was associated with GI symptoms, suggesting these patients may experience dysregulation of the brain-gut axis, which has been associated with elevated faecal SCG3 in patients with irritable bowel syndrome.^36^

Cognitive impairment was associated with elevated Neurofascin (NFASC OR=1.05), Spondin-1 (OR=1.03) and Iduronate sulfatase (IDS OR=1.04; fig 1F&K). NFASC and Spondin-1 regulate neural growth and NFASC was associated with the highest odds of this outcome.^37, 38^ IDS is an X-linked lysosomal enzyme which degrades ECM proteoglycans to enable leucocyte infiltration into tissues.^39, 40^ These findings suggest neuroinflammation and a resultant tissue repair response, supported by our findings that elevated C1QA (OR=1.04), DPP10 (OR=1.03) and CCL11 (OR=1.02) were also associated with cognitive impairment.

To validate the findings from PLR analysis, we examined the distribution of data, prioritising proteins that were most strongly predictive of each symptom outcome. Each protein in question was significantly elevated in the symptom group compared to recovered (fig 1G–K & S1), confirming that the observed protein signatures differentiate adults with long COVID. However, the marked overlap indicates these markers are not useful on an individual basis as biomarkers.

We next sought to understand inflammatory responses in patients with cardiorespiratory symptoms, the largest group in our analysis. In addition to sCD58, elevated markers of tissue repair, including Delta/notch-like EGF repeat (DNER OR=0.82) and IDS (OR=0.78) were associated with reduced risk of cardiorespiratory symptoms (fig 1B & 2A). DNER was also protective for all groups except GI (fig 1B–F) and is known to enhance cell proliferation and differentiation.^41, 42^ Notably, elevated IDS associated with recovery compared to cardiorespiratory symptoms (OR=0.78) and all other symptom groups, except cognitive impairment where IDS predicted this outcome (fig 1B–F).

**Figure 2.**
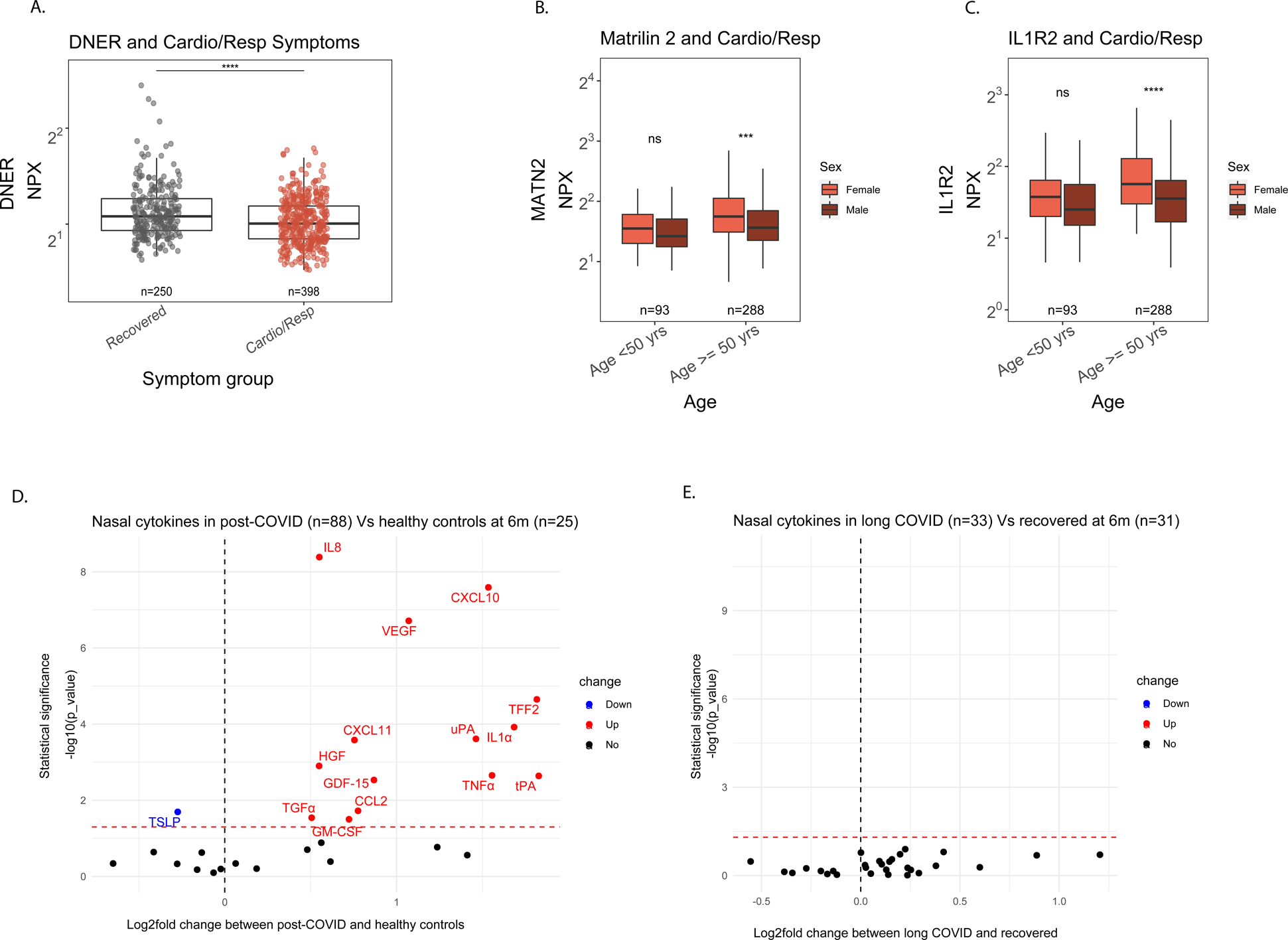
Local and systemic inflammatory profiles in patients with cardiorespiratory symptoms. DNER levels between recovered patients and those with cardiorespiratory symptoms (A). Patients with cardiorespiratory symptoms were divided by age group and sex to assess whether differences in oestrogen levels between men and women influence inflammatory protein levels in women with cardiorespiratory symptoms (B & C). Volcano plots showing the log_2_-fold change in nasal cytokine concentration between 88 post-COVID patients and 25 pre-pandemic healthy control samples (D) and between long COVID patients (n=33) of which 29 experienced respiratory symptoms, and 31 post-COVID patients who felt recovered (E). The red values indicate increased cytokine levels between groups and a statistically significant change (*p*<0.05). The blue values denote depressed cytokine levels and a statistically significant change (*p*<0.05). Median values were compared between groups using the Wilcoxon signed-rank test after FDR adjustment. * = p<0·05, ** = p<0·01, *** = p<0·001, ****= p<0·0001.

IDS maintains tissues by preventing accumulation of ECM proteoglycans and facilitating leucocyte migration into tissues.^39, 40^ IDS, may have divergent functions in different tissue environments, for example supporting lung tissue repair and preventing post-COVID breathlessness, whilst encouraging neuroinflammation and thus cognitive impairment. Our data suggests immunosuppressive factors and a robust tissue repair response may prevent cardiorespiratory symptoms after COVID-19.

Consistent with previous reports,^11^ we found women were more likely to experience long COVID symptoms (Fig 1B–F; table 1). Since oestrogen is known to influence immunological responses,^43^ we compared protein levels between men and women younger and older than 50 years. Inflammation markers IL1R2 and MATN2 were significantly higher in post-menopausal women (> 50 years) with cardiorespiratory symptoms (IL1R2 p=0.0002; MATN2 p<0.0001; fig 2B&C). Similar trends were seen in fatigue and anxiety/depression (figure S2A–C). Oestrogen-dependent differences would be expected to be most pronounced in pre-menopausal women,^44^ but we found no evidence of this. There is evidence women have stronger innate immune responses to infection^43, 45^ and are known to be at greater risk of autoimmunity for several reasons.^43^ These mechanisms could explain the observed enhancement of monocytic inflammation and complement activation in women with cardiorespiratory symptoms, fatigue and anxiety/depression.

Examining proteins associated with GI symptoms, there were no significant differences seen between men and women (fig S2D–F). In the cognitive impairment group IDS was significantly higher in pre-menopausal women (*p*=0.02), though this effect was lost in the post-menopausal group (fig S2G). *IDS* is X-linked, which may in part explain these differences. ^46^ Thus, while non-hormonal differences in immune responses between men and women may explain the increased likelihood of women to experience some long COVID symptoms, variations in X-linked silencing of the *IDS* gene may in part explain sex differences seen in post-COVID cognitive impairment.

We considered whether SARS-CoV-2 persistence might drive systemic inflammation.^47, 48^ We performed an exploratory analysis of sputum from 23 adults with long COVID, who all reported cardiorespiratory symptoms, and 17 recovered adults 6 months after hospitalisation. Broncho-alveolar lavage fluid from SARS-CoV-2 naïve individuals were used as controls. Although low concentrations of N antigen were detected in 4 samples, there was no difference between those with symptoms and those who were fully recovered (fig S3A); furthermore, S antigen was undetectable in all sputum samples. These findings do not exclude viral persistence and the underlying cause of ongoing inflammation remains uncertain.

To determine the source of systemic inflammation in patients with cardiorespiratory symptoms, we analysed nasosorption samples from 88 adults 6 months after hospitalisation compared to 25 healthy controls (table S2). Several inflammatory markers were significantly elevated in post-COVID adults (fig 2D); in addition to TNFα, IL8 and IL-1α, several epithelial growth factors were elevated including VEGF, TGFα, GDF and HGF. However, there was no difference in nasal mediator levels between those recovered (n=31) and those who were not (n=33) (fig 2E). In adults with cardiorespiratory symptoms (n=29), inflammatory mediators elevated in plasma were not elevated in the upper respiratory tract (fig S3B-G). Furthermore, in those who had matched plasma and nasal inflammatory mediator measurements (n=70) there was no correlation between mediator levels at different sites (fig S3H–M). This exploratory analysis suggests protein levels in plasma poorly reflect inflammation occurring in specific tissue sites, and that inflammation in the upper respiratory tract is not associated with post-COVID breathlessness. We did not measure inflammation in the lower respiratory tract, which might contribute to post-COVID breathlessness.^23^

Our study has important limitations. It was not designed for biomarker discovery. The use of PLR to analyse high-dimensional data revealed subtle differences in protein levels that are not striking when viewed unidimensionally (fig 1G–K & S1). Univariate analyses do not account for the combined and adjusted effects of all explanatory variables. PLR analysis is better suited to high dimensional data accounting for collinearity and avoiding false discovery.

In conclusion, in a large, hospitalised cohort we found indications of monocytic inflammation and complement activation in those with continued symptoms 6 months after COVID-19 (fig 3). We found distinctive inflammatory patterns in those with cognitive impairment and GI symptoms. These findings add to evidence that prolonged symptoms arise from different causes and that therapeutic trials need to take this diversity into account.

**Figure 3.**
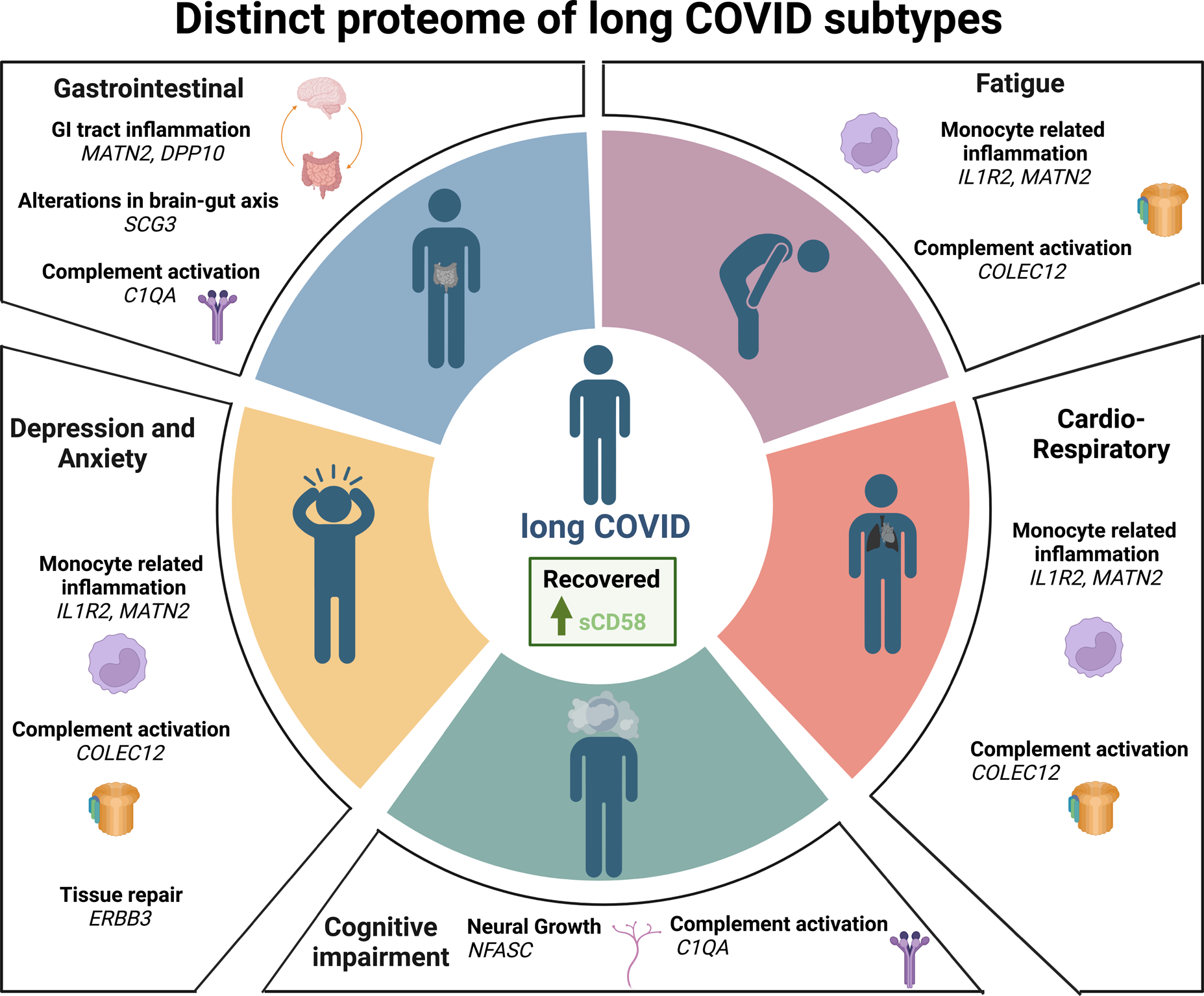
Graphical Abstract. Protein signatures associated with each long COVID subtype are shown. The blood proteome of 719 patients was analysed, 6 months after COVID-19 hospitalisation. For all markers shown, elevated levels were associated with each symptom outcome. However, elevated sCD58 was associated with feeling recovered.

## Methods (Online)

### Study design and Ethics

After hospital discharge patients >18 years old who had no co-morbidity resulting in a prognosis of less than 6 months, who had been hospitalised for COVID-19 were recruited to the PHOSP-COVID study. Patients that had been hospitalised between February 2020 and January 2021 were recruited. Both sexes were recruited and gender was self-reported. Written informed consent was obtained from all patients. Ethical approvals for the PHOSP-COVID study were given by Leeds West Research Ethics Committee (20/YH/0225).

Clinical data and plasma samples were prospectively collected from adult cases of COVID-19 approximately 6 months after hospitalisation, via the PHOSP-COVID multicentre UK study.^11^ Data relating to patient demographics and the acute admission were collected via the ISARIC4C study.^49^ Adults hospitalised during the SARS-CoV-2 pandemic were systematically recruited into the International Severe Acute Respiratory and Emerging Infection Consortium (ISARIC) World Health Organization Clinical Characterisation Protocol UK (IRAS260007 and IRAS126600).^49^ Written informed consent was obtained from all patients. Ethical approval was given by the South Central– Oxford C Research Ethics Committee in England (reference: 13/SC/0149), Scotland A Research Ethics Committee (20/SS/0028) and World Health Organization Ethics Review Committee (RPC571 and RPC572l; 25 April 2013).

Data were collected to account for variables which may affect symptom outcome, via hospital records and self-reporting. Disease severity was classified according to the WHO Clinical Progression score.^50^ Clinical data were used to place patients into 6 categories: ‘Recovered’, ‘GI’, ‘Cardiorespiratory’, ‘Fatigue’, ‘Cognitive impairment’ and ‘Anxiety/depression’ (Table S1). Patient reported symptoms and validated clinical scores were used including: MRC breathlessness score, dyspnoea-12 score, FACIT score, PHQ-9 and GAD-7. Responses to symptom questionnaires about chest pain and palpitations were also used. Cognitive impairment was defined as a Montreal Cognitive Assessment (MoCA) score <26. GI symptoms were defined as answering ‘Yes’ to the presence of at least two out of the following: abdominal pain, nausea and/or vomiting, weight loss, diarrhoea, constipation. ‘Recovered’ was defined by self-reporting and/or below threshold scores from the clinical questionnaires. Patients were placed in multiple groups if they experienced a combination of symptoms (fig 1A).

Matched nasal fluid and sputum samples were collected from a subgroup of convalescent patients approximately 6 months after hospitalisation via the PHOSP-COVID study. Nasal fluid collected from healthy volunteers prior to the COVID-19 pandemic and bronchoalveolar lavage fluid (BALF) collected from SARS-COV-2 naïve individuals were used as controls (Table S2; fig 2D & S3A). Written consent was obtained for all individuals and ethical approvals were given by London-Harrow Research Ethics Committee (13/LO/1899) for the collection of nasal samples and the Health Research Authority London–Fulham Research Ethics Committee (IRAS Project ID 154109; references 14/LO/1023, 10/H0711/94, and 11/LO/1826) for BALF samples.

### Procedures

EDTA plasma was collected from whole blood taken by venepuncture and frozen at −80°C as previously described.^8^ Nasal fluid was collected using a NasosorptionTM FX·I device (Hunt Developments UK Ltd), which uses a synthetic absorptive matrix to collect concentrated nasal fluid. Samples were stored and eluted as previously described.^51^ Sputum samples were collected via passive expectoration and frozen at −80°C without the addition of buffers. Sputum samples taken from convalescent patients were compared to BALF collected from healthy SARS-CoV-2 naïve controls. BALF samples were used to act as a comparison for lower respiratory tract samples since passively expectorated sputum from healthy SARS-CoV-2 naïve individuals is challenging to obtain. BALF samples were obtained by instillation and recovery of up to 240 ml of normal saline via a fiberoptic bronchoscope. BALF was filtered through 100µM strainers into sterile 50ml Falcon tubes, then centrifuged for 10 minutes at 400g at 4°C. The resulting supernatant was transferred into sterile 50ml Falcon tubes and frozen at −80°C until use. The full methods for BALF collection and processing have been described.^52, 53^

### Immunoassays

To determine inflammatory signatures which associated with symptom outcomes, plasma samples were analysed on an Olink Explore 384 Inflammation panel (Uppsala, Sweden)^9^. Data were first normalized to an extension control that was included in each sample well. Plates were standardized by normalizing to inter-plate controls run in triplicate on each plate. All samples were run in a single batch to prevent batch-effects. Final normalized relative protein quantities were reported as log2 normalized protein expression (NPX) values.

Sputum samples were thawed prior to analysis and sputum plugs were extracted with the addition of 0.1% DTT creating a 1 in 2 sample dilution, as previously described.^54^ SARS-CoV-2 Spike and Nucleocapsid proteins were measured by electrochemiluminescence S-plex assay at a fixed dilution of 1 in 2 (Mesoscale Diagnostics, Rockville, Maryland, USA), as per the manufacturers protocol.^55^ Control BALF samples were thawed and measured on the same plate, neat. The S-plex assay is highly sensitive in detecting viral antigen in respiratory tract samples.^56^

Nasal cytokines were measured by electrochemiluminescence (MSD) and Luminex bead multiplex assays (Biotechne, Minneapolis, United States). The V-plex pro-inflammatory 1 kit (MSD), R-plex custom kit (MSD) and Human Premixed Multi-analyte custom kit (Biotechne) were used. Nasal samples were analysed at a fixed dilution of 1 in 2 using the R-plex and Luminex assays, and neat using the proinflammatory panel 1 kit. MSD plates were measured on a MESO QuickPlex SQ 120 Reader (MSD) and Luminex plates on a BioPlex200 instrument (Bio-Rad, UK). Cytokine concentrations were calculated using a reference standard and assigned pg/mL. All values at or below the lower limit of detection (LLOD) were replaced with LLOD. All values at or above the upper limit of detection (ULOD) were replaced with ULOD. The full methods and list of analytes are detailed in the supplements.

### Statistics

To determine protein signatures which predicted each symptom outcome, a ridge penalised logistic regression (PLR) was used. PLR shrinks coefficients to account for high dimensional data and multicollinearity given the large number of predictor variables measured. The model alpha value was set at zero to capture the coefficients of all proteins measured, given that this was an exploratory analysis with no prior selection of putative mediators. A 50 repeats 10-fold nested cross-validation was used to select the models with lowest classification error as well as the optimal lambda for each model. To increase the sensitivity and specificity of the PLR comparing Cognitive impairment (n=65) and Recovered (n=250) groups, the model was weighted to account for the imbalance in group size and prevent bias towards cases being classified as the majority class (Recovered). The weighted model had a sensitivity of 0.98. The distribution of classification error and AUC generated by the cross-validation is shown in figure S4.

Protein measurements, age, sex, acute disease severity and pre-existing comorbidities were included as covariates (table S3). Ethnicity was not included as a covariate since ethnicity has been previously shown not to predict symptom outcome in this cohort.^11^ Individuals with missing covariate data were excluded from the regression analysis. Each symptom group was compared to patients from the ‘Recovered’ group as the model’s binary outcome. The model coefficients of each covariate were converted into odds ratios for each outcome and visualised in a forest plot, after removing variables associated with adjusted odds ratios between 0.09 and 1.02, since these are unlikely to be of clinical significance using the ridge penalisation method which shrinks coefficients according to the relative importance of the variable. Since confidence intervals cannot appropriately be derived from penalised models, error bars were calculated using the median accuracy of the model generated by the nested cross-validation (fig S4). Analyses were performed in R version 4.2.0 using *‘lme4’, ‘caret’, ‘glmnet’* and *‘ggplot2’* packages. The outputs from the model were validated in a univariate analysis using Wilcoxon signed rank test and FDR adjustment.

To determine if differences in protein levels between men and women related to hormonal differences, we divided each symptom group into pre-menopausal and post-menopausal groups using an age cut-off of 50 years old. Differences between sexes in each group were determined using the Wilcoxon-signed rank test. To understand if antigen persistence contributed to inflammation in adults with long COVID, the median viral antigen concentration from sputum/BALF samples and cytokine concentrations from nasal samples were compared using the Wilcoxon signed rank test. All tests were two-tailed and statistical significance was defined as a *p*-value<0.05 after adjustment for false discovery rate (q-value=0.05). Analyses were performed in R version 4.2.0 using *‘ggpubr’* and *‘ggplot2’* packages.

## Supporting information

ISARIC4C and PHOSP-COVID

ISARIC4C and PHOSP-COVID

## Data Availability

This is an Open Access article under the CC BY 4.0 license. 
The PHOSP-COVID protocol, consent form, definition and derivation of clinical characteristics and outcomes, training materials, regulatory documents, information about requests for data access, and other relevant study materials are available online: https://phosp.org/resource/. Access to these materials can be granted by contacting.
The ISARIC4C protocol, data sharing and publication policy are available at https://isaric4c.net. The ISARIC4C Independent Data and Material Access Committee welcomes applications for access to data and materials (https://isaric4c.net).
All data used in this study is available within ODAP and accessible under reasonable request. Data access criteria and information about how to request access is available online: https://phosp.org/resource/. If criteria are met and a request is made, access can be gained by signing the eDRIS user agreement.

## Data sharing

This is an Open Access article under the CC BY 4.0 license.

The PHOSP-COVID protocol, consent form, definition and derivation of clinical characteristics and outcomes, training materials, regulatory documents, information about requests for data access, and other relevant study materials are available online: https://phosp.org/resource/. Access to these materials can be granted by contacting.

The ISARIC4C protocol, data sharing and publication policy are available at https://isaric4c.net. ISARIC4C’s Independent Data and Material Access Committee welcomes applications for access to data and materials (https://isaric4c.net).

All data used in this study is available within ODAP and accessible under reasonable request. Data access criteria and information about how to request access is available online: https://phosp.org/resource/. If criteria are met and a request is made, access can be gained by signing the eDRIS user agreement.

## Acknowledgements

This research used data assets made available by Outbreak Data Analysis Platform (ODAP) as part of the Data and Connectivity National Core Study, led by Health Data Research UK in partnership with the Office for National Statistics and funded by UK Research and Innovation (grant ref MC_PC_20058).

This work is supported by the following grants: The PHOSP-COVD study is jointly funded by UK Research and Innovation and National Institute of Health and Care Research (grant references: MR/V027859/1 and COV0319). ISARIC4C is supported by grants from the National Institute for Health and Care Research (award CO-CIN-01) and the Medical Research Council (grant MC_PC_19059) Liverpool Experimental Cancer Medicine Centre provided infrastructure support for this research (grant reference: C18616/A25153). Other grants which have supported this work include: the UK Coronavirus Immunology Consortium [funder reference:1257927], the Imperial Biomedical Research Centre (NIHR Imperial BRC, grant IS-BRC-1215-20013), the Health Protection Research Unit (HPRU) in Respiratory Infections at Imperial College London and NIHR HPRU in Emerging and Zoonotic Infections at University of Liverpool, both in partnership with Public Health England, [NIHR award 200907], Wellcome Trust and Department for International Development [215091/Z/18/Z], Health Data Research UK (HDR UK) [grant code: 2021.0155], Medical Research Council [grant code: MC_UU_12014/12], and NIHR Clinical Research Network for providing infrastructure support for this research.

FL is supported by an MRC clinical training fellowship [award MR/W000970/1]. CE is funded by NIHR [grant P91258-4]. LPH is supported by Oxford NIHR Biomedical Research Centre. AART is supported by a BHF Intermediate Clinical Fellowship (FS/18/13/33281). SLRJ receives support from UKRI, GCRF, Rosetrees Trust, BHIVA, EDCTP, Globvac. JDC has grants from AstraZeneca, Boehringer Ingelheim, GlaxoSmithKline, Gilead Sciences, Grifols, Novartis and Insmed. RAE holds a NIHR Clinician Scientist Fellowship (CS-2016-16-020). AH is currently supported by UKLJResearch and Innovation. NIHR and NIHR Manchester BRC. BR receives support from BHF Oxford Centre of Research Excellence, NIHR Oxford BRC and MRC. DGW is supported by an NIHR Advanced Fellowship. AH has received support from MRC and for the Coronavirus Immunology Consortium (<u>MR/V028448/1</u>). LVW has received support from UKRI, GSK/Asthma + Lung UK and NIHR for this study. MGS has received support from NIHR UK, MRC UK and Health Protection Research Unit in Emerging & Zoonotic Infections, University of Liverpool. JKB is supported by the Wellcome Trust (223164/Z/21/Z) and UKRI (MC_PC_20004, MC_PC_19025, MC_PC_1905, MRNO2995X/1, and MC_PC_20029). LT is supported by the Wellcome Trust [clinical career development fellowship grant number 205228/Z/16/Z], the Centre of Excellence in Infectious Diseases Research (CEIDR) and the Alder Hey Charity. PJMO is supported by a NIHR Senior Investigator Award [award 201385].

The funders were not involved in the study design, interpretation of data or writing of this manuscript. The views expressed are those of the authors and not necessarily those of the DHSC, DID, NIHR, MRC, the Wellcome Trust, UK-HAS, the National Health Service, or the Department of Health.

This study would not be possible without all the participants who have given their time and support. We thank all the participants and their families. We thank the many research administrators, health-care and social-care professionals who contributed to setting up and delivering the PHOSP-COVID study at all of the 65 NHS trusts/Health boards and 25 research institutions across the UK, as well as those who contributed to setting up and delivering the ISARIC4C study at 305 NHS trusts/ Health boards. We also thank all the supporting staff at the NIHR Clinical Research Network, Health Research Authority, Research Ethics Committee, Department of Health and Social Care, Public Health Scotland, and Public Health England. We thank Kate Holmes at the NIHR Office for Clinical Research Infrastructure (NOCRI) for her support in coordinating the charities group. The PHOSP-COVID industry framework was formed to provide advice and support in commercial discussions, and we thank the Association of the British Pharmaceutical Industry as well NOCRI for coordinating this. We are very grateful to all the charities that have provided insight to the study: Action Pulmonary Fibrosis, Alzheimer’s Research UK, Asthma + Lung UK, British Heart Foundation, Diabetes UK, Cystic Fibrosis Trust, Kidney Research UK, MQ Mental Health, Muscular Dystrophy UK, Stroke Association Blood Cancer UK, McPin Foundations, and Versus Arthritis. We thank the NIHR Leicester Biomedical Research Centre patient and public involvement group and Long Covid Support. We would also like to thank Professor Golam Khandaker and Professor Dawn C. Newcomb who provided valuable feedback on this work. Figure 3 was created using Biorender.

## Contributors

**FL** recruited participants, acquired clinical samples, analysed and interpreted data and co-wrote the manuscript, including all drafting and revisions. **CE** analysed and interpreted data and co-wrote this manuscript, including all drafting and revisions**. SF** and **MR** supported analysis and interpretation of data as well as drafting and revisions. **DS, JKS, SCM, SA, NM, JN, CK, OCL, OE, HJCM, ASh, ASi, MS, VCH, MT, NJG, NIL, CC** contributed to acquisition of data underlying this study. **LHW**, **AART, SLRJ, LH, OMK, DGW, TIdS** and **AH** made substantial contributions to conception/design and implementation of this work and/or acquisition of clinical samples for this work. They have supported drafting and revisions of the manuscript. **EH, JKQ** and **ABD** made substantial contributions to study design as well as data access, linkage and analysis. They have supported drafting and revisions of this work. **JDC, LPH, AH, BR, KP, MM, WG** made substantial contributions to conception and design of this work and have supported drafting and revisions of this work. **JKB** obtained funding for ISARIC4C, is ISARIC4C consortium co-lead, has made substantial contributions to conception and design of this work and has supported drafting and revisions of this work. **MGS** obtained funding for ISARIC4C, is ISARIC4C consortium co-lead, sponsor/protocol chief investigator, has made substantial contributions to conception and design of this work and has supported drafting and revisions of this work. **RAE and LVW** are co-leads of PHOSP-COVID, made substantial contributions to conception and design of this work, acquisition and analysis of data, and have supported drafting and revisions of this work. **CB** is the chief investigator of PHOSP-COVID and has made substantial contributions to conception and design of this work. **RST** and **LT** made substantial contributions to acquisition, analysis and interpretation of the data underlying this study and have contributed to drafting and revisions of this work. **PJMO** obtained funding for ISARIC4C, is ISARIC4C consortium co-lead, sponsor/protocol chief investigator, and has made substantial contributions to conception and design of this work. **RST** and **PJMO** have also made key contributions to interpretation of data and have co-written this manuscript. All authors have read and approve the final version to be published. All authors agree to accountability for all aspects of this work.

All investigators within ISARIC4C and the PHOSP-COVID consortia have made substantial contributions to the conception or design of this study and/or acquisition of data for this study. The full list of authors within these groups is available in the supplementary materials.

---

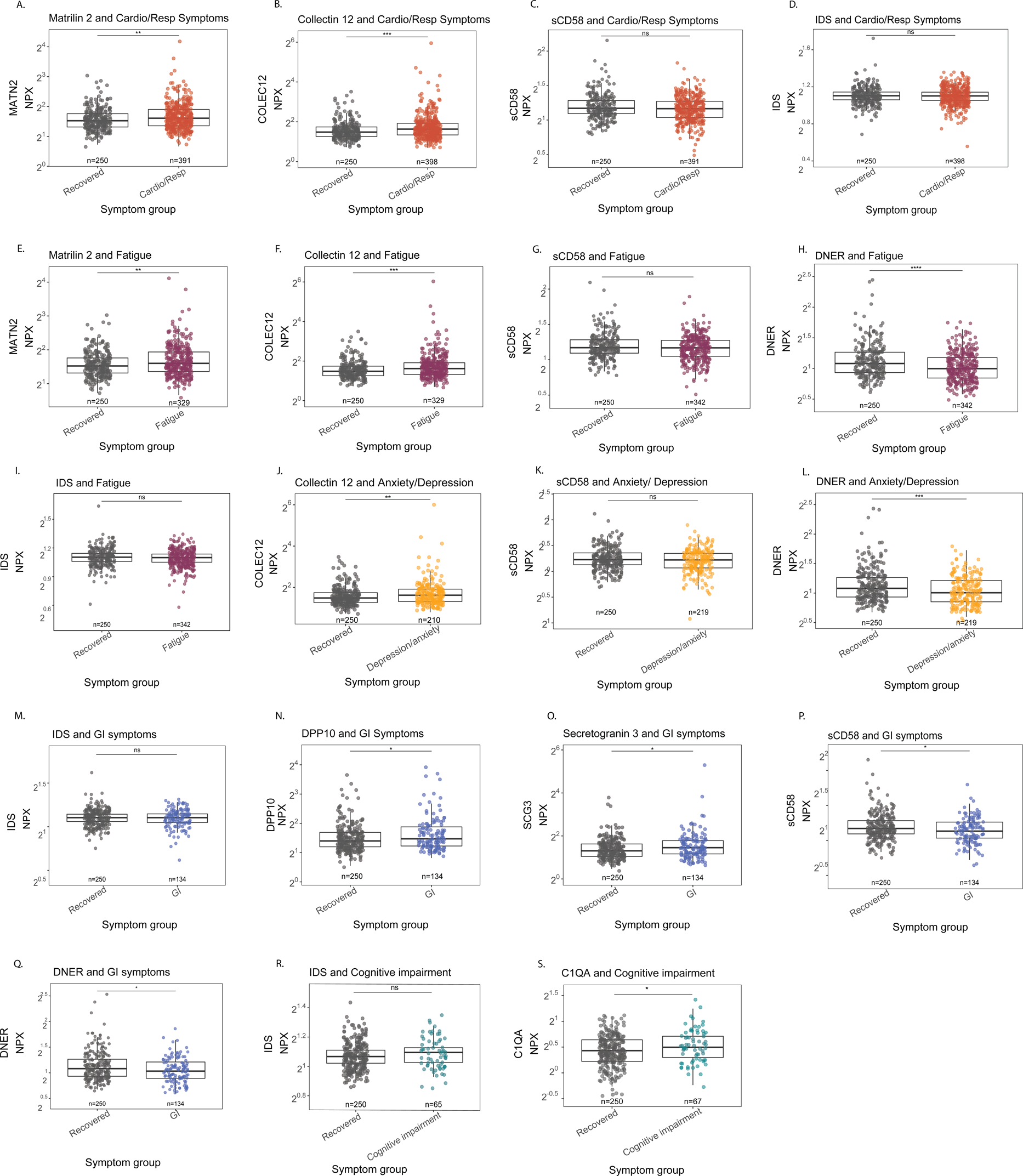
Figure S1.

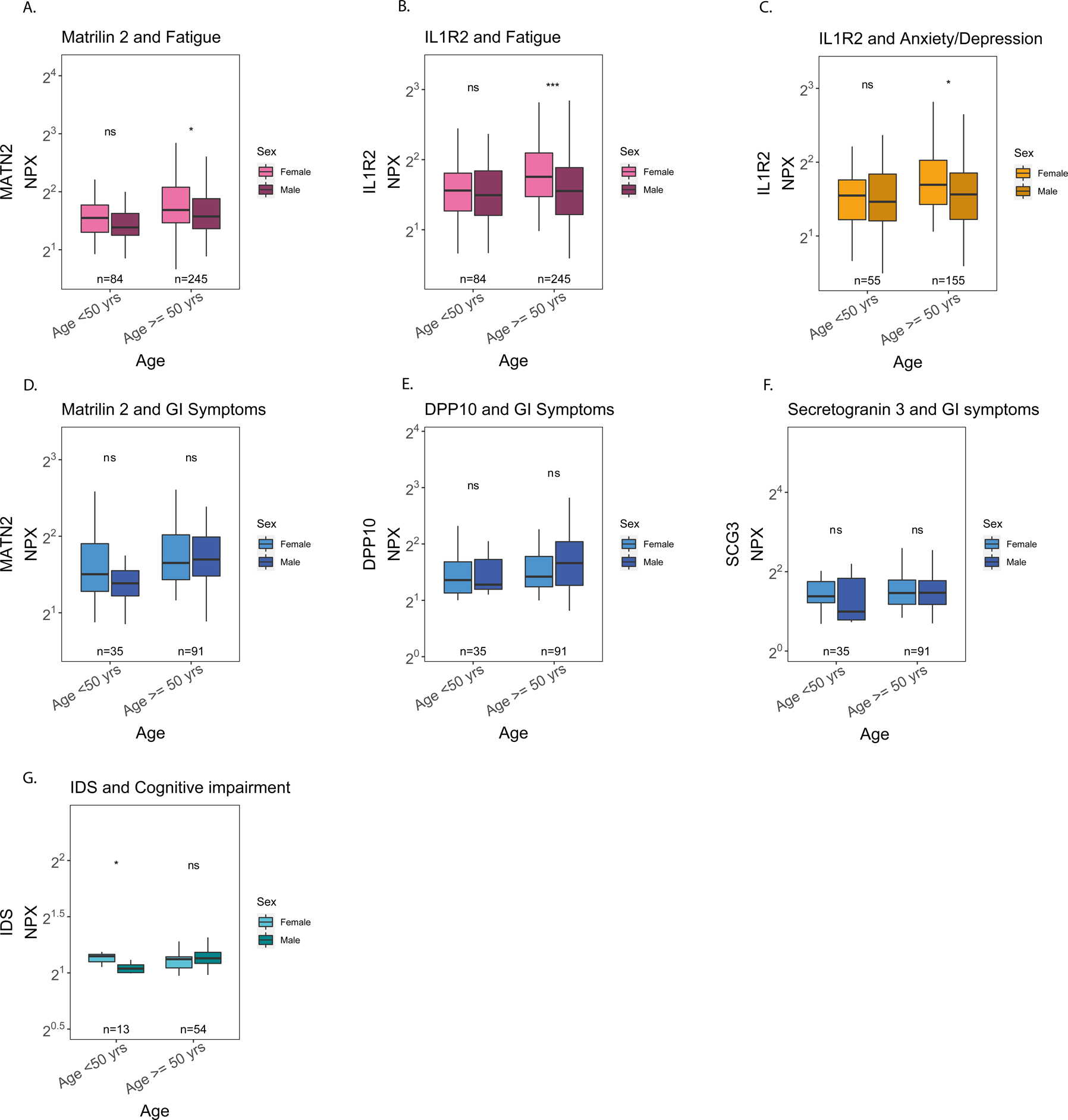
Figure S2.

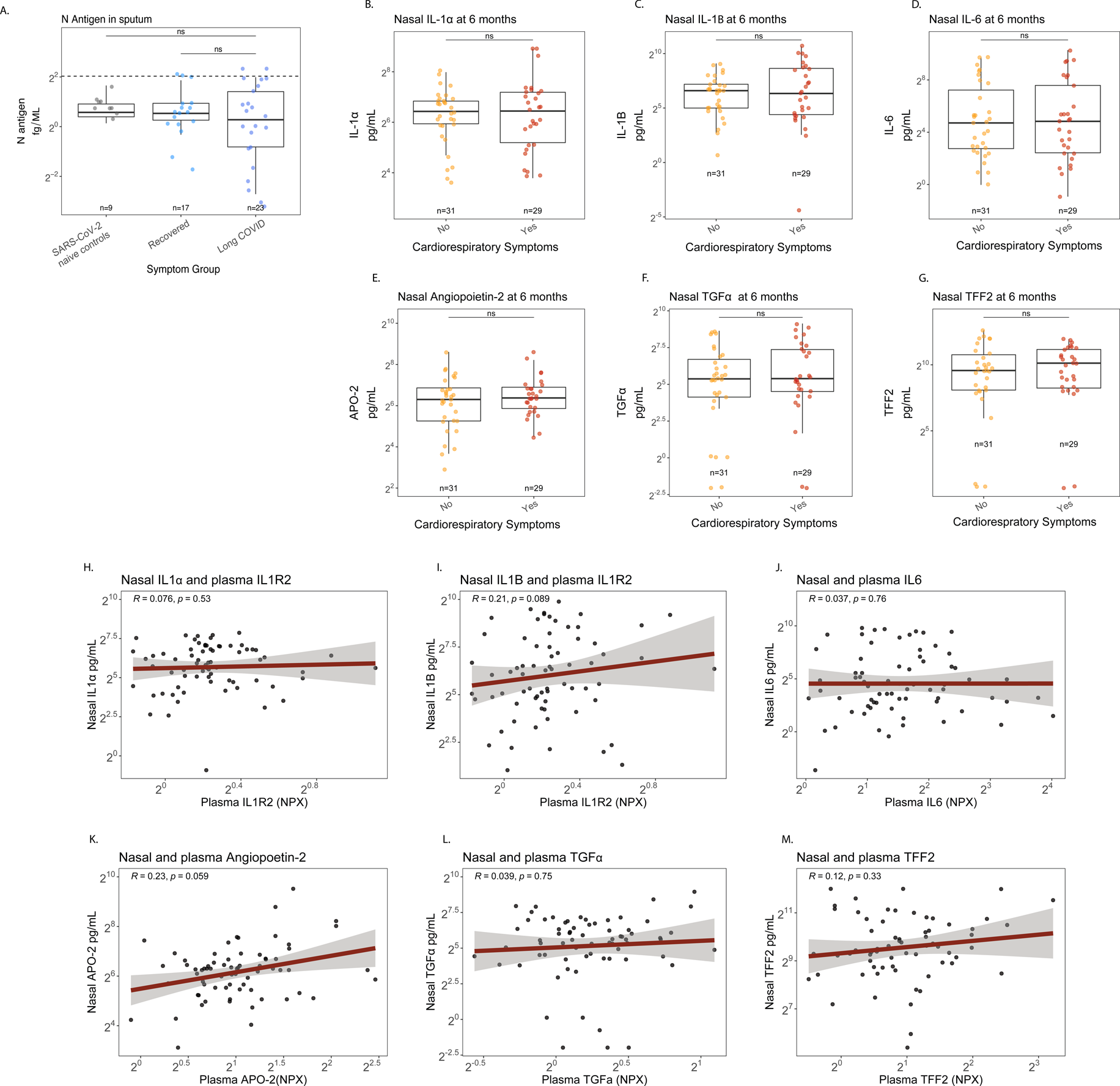
Figure S3.

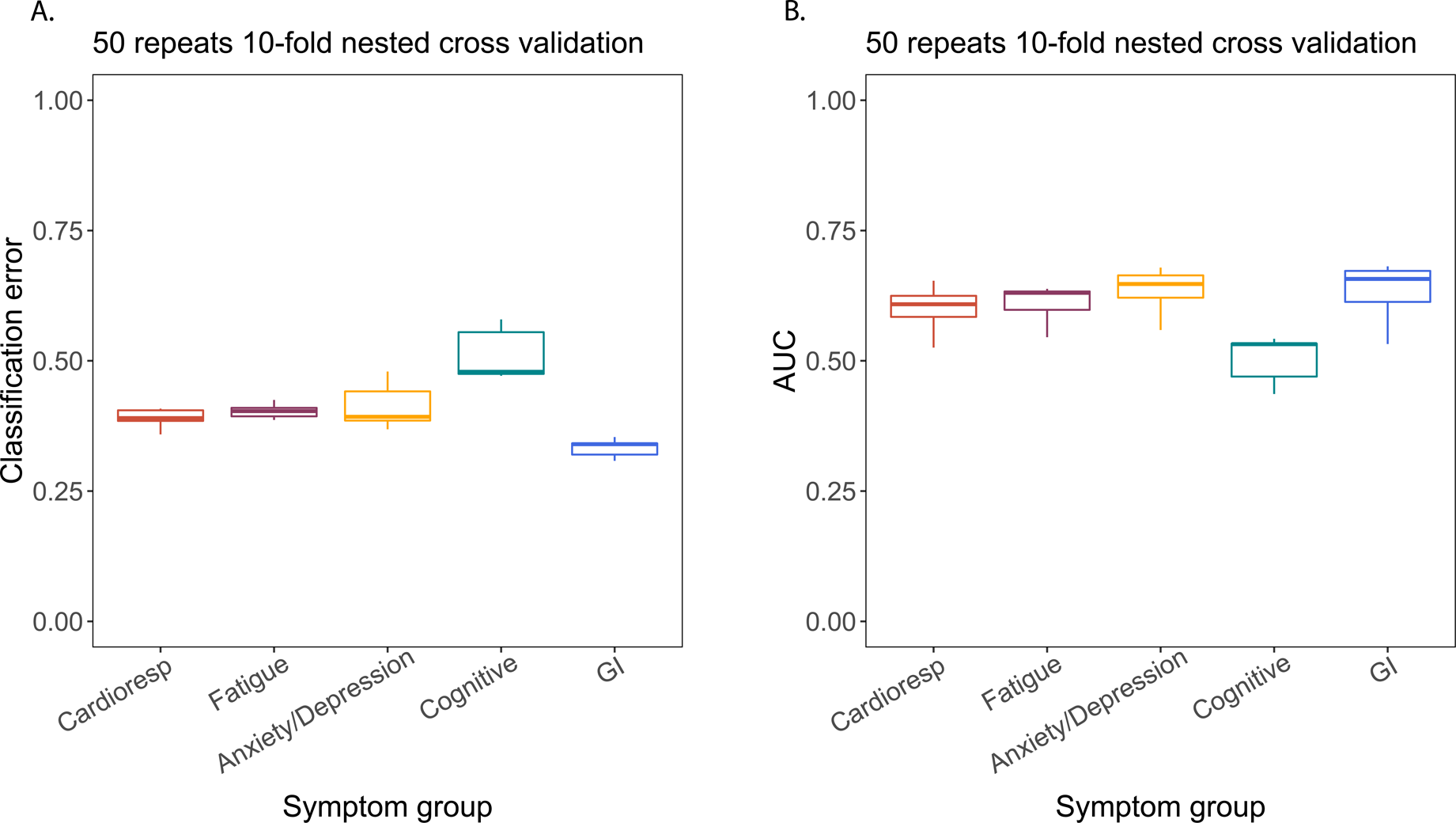
Figure S4.

## Notes

### Competing Interest Statement

FL, CE, DS, JKS, SCM, CD, CK, NM, LN, EMH, ABD, JKQ, LPH, KP, LH, OMK, SF, TIdS, DGW, RST and JKB have no conflicts of interest. AART receives speaker fees and support to attend meetings from Janssen Pharmaceuticals. SLRJ is on the data safety monitoring board for Bexero trial in HIV+ adults in Kenya. JDC is the deputy chief editor of ERS and receives consulting fees from AstraZeneca, Boehringer Ingelheim, Chiesi, GlaxoSmithKline, Insmed, Janssen, Novartis, Pfizer and Zambon. AH is Deputy chair of NIHR Translational Research Collaboration (unpaid role). BR receives honoraria from Axcella therapeutics. RAE is co-lead of PHOSP-COVID and receives fees from Astrazenaca / Evidera for consultancy on Long Covid and from Astrazenaca for consultancy on digital health. RAE has received speaker fees from Boehringer in June 2021 and has held a role as European Respiratory Society Assembly 01.02 Pulmonary Rehabilitation secretary. RAE is on the American Thoracic Society Pulmonary Rehabilitation Assembly programme committee. LVW also receives funding from Orion pharma and GSK and holds contracts with Genentech and AstraZenaca. LVW has received consulting fees from Galapagos and Boehringer, is on the data advisory board for Galapagos and is Associate Editor for European Respiratory Journal. AH is a member of NIHR Urgent Public Health Group (June 2020-March 2021). MM is an applicant on the PHOSP study funded by NIHR/DHSC. MGS acts as an independent external and non-remunerated member of Pfizers External Data Monitoring Committee for their mRNA vaccine program(s), is Chair of Infectious Disease Scientific Advisory Board of Integrum Scientific LLC, Greensboro, NC, USA and is director of MedEx Solutions Ltd and majority owner of MedEx Solutions Ltd and minority owner of Integrum Scientific LLC, Greensboro, NC, USA. MGS has been in receipt of gifts from Chiesi Farmaceutici S.p.A. of Clinical Trial Investigational Medicinal Product without encumbrance and distribution of same to trial sites. MGS is a non-renumerated member of HMG UK New Emerging Respiratory Virus Threats Advisory Group (NERVTAG) and has previously been a non-renumerated member of SAGE. CB has received consulting fees and/or grants from GSK, AZ, Genentech, Roche, Novartis, Sanofi, Regeneron, Chiesi, Mologic and 4DPharma. LT has received consulting fees from MHRA and speak fees from Eisai Ltd. LT has a patent pending with ZikaVac. PJMO reports grants from the EU Innovative Medicines Initiative (IMI) 2 Joint Undertaking during the submitted work; grants from UK Medical Research Council, GlaxoSmithKline, Wellcome Trust, EU-IMI, UK, National Institute for Health Research, and UK Research and Innovation-Department for Business, Energy and Industrial Strategy; and personal fees from Pfizer, Janssen, and Seqirus, outside the submitted work.

### Clinical Protocols

https://phosp.org/resource/

### Author Declarations

This study uses data from the PHOSPCOVID and ISARIC4C study. Written informed consent was obtained from all patients. Ethical approvals for the PHOSPCOVID study were given by Leeds West Research Ethics Committee (20/YH/0225). Ethical approval was given by the South Central-Oxford C Research Ethics Committee in England (reference: 13/SC/0149), Scotland A Research Ethics Committee (20/SS/0028) and World Health Organization Ethics Review Committee (RPC571 and RPC572l; 25 April 2013).

