## supplements; supplementary material; table S1; table S2; y=table S3 for "Large scale phenotyping of long COVID inflammation reveals mechanistic subtypes of disease"

**Supplemental materials**

**Supplemental methods**

*Nasal cytokine immunoassay*

Nasal Angiopoeitin-2, CXCL-10, Galectin-9, GM-CSF, IL-1α, IL-33, TSLP, CXCL11, VEGF, CCL2, CXCL11, GDF-15, HGF, Thrombomodulin and IL-15 were measured by custom Luminex discovery bead-based ELISA (Biotechne, Minneapolis, United States). IFNγ, IL-1β, IL-2, IL-4, IL-6, IL-8, IL-19, IL-12p70, IL-13 and TNFα were measured using the Proinflammatory Panel 1 V-plex kit, which uses an MSD electrochemiluminescence multiplex assay (Mesoscale Diagnostics, Rockville, Maryland, USA). Trefoil factor 2 (TFF2), Tissue Plasminogen Activator (tPA), Follistatin and TGFa were measured using an R-plex custom electrochemiluminescence multiplex assay (MSD).

Nasal samples were thawed at room temperature and analysed at a fixed dilution of 1 in 2 using the R-plex and Luminex assays, and neat using the Proinflammatory Panel 1 V-plex kit. Plates were prepared and analysed according to the manufacturers protocol.^1–3^ MSD V-plex plates consisted of 96 wells with 10 spots, each precoated with a capture antibody for the given analyte, and to which neat sample was added and incubated. The R-plex custom plate consisted of 96 wells with 4 spots coated with streptavidin. A solution of biotinylated capture antibody for each analyte was incubated on each R-plex plate to enable measurement of the chosen analytes. Nasal fluid was then added and incubated, followed by the addition of a labelled detection antibody. MSD plates were measured on a MESO QuickPlex SQ 120 Reader (MSD) and cytokine concentrations were calculated using a reference standard and assigned pg/ml. For the Luminex assay, samples were incubated on a 96 well plate with addition of beads labelled with the analyte capture antibody. Luminex plates were read on a BioPlex200 instrument (Bio-Rad, UK) and cytokine concentrations were derived from the reference standard and assigned pg/ml. All values at or below the lower limit of detection (LLOD) were replaced with LLOD. All values at or above the upper limit of detection (ULOD) were replaced with ULOD.

*Viral antigen measurements from sputum*

Sputum samples were thawed at room temperature and sputum plugs were extracted and weighed. Eight millilitres of PBS were added per mg of sputum to create a solution and samples were centrifuged at 400g for 10 minutes at 4°C. Four volumes of PBS supernatant were then removed and stored at -80°C. Another 4 volumes were removed for the addition of 4 volumes of 0.1% dithiothreitol (DTT), creating a 1 in 2 dilution, and stored at -80°C. DTT was added to reduce matrix effects and enable measurement of sputum proteins, and this method has been previously validated.^4^ Sputum diluted in DTT was used for the measurement of viral Spike (S) and Nucleopcapsid (N) antigens using a MSD S plex SARS-CoV-2 N kit and SARS-CoV-2 S kit (MSD). The kits used an electrochemiluminescence assay and plates consisted of 96 wells containing 1 spot coated with a streptavidin bound biotinylated capture antibody. Plates were blocked with MSD Blocker A, followed by the addition of sputum samples in DTT, analysed at a fixed dilution of 1 in 2. After sample incubation a TURBO-BOOST detection antibody was added followed by the addition of a TURBO-TAG which enables higher sensitivity of the assay.^5^ All values at or below the lower limit of detection (LLOD) were replaced with LLOD. All values at or above the upper limit of detection (ULOD) were replaced with ULOD.

**Supplementary tables**

| **Symptom group** | **Questionnaire** | **Response type** | **Threshold score** |
| --- | --- | --- | --- |
| Cardiorespiratory* | MRC dyspnoea scale | Numerical scale | Score >1 |
|  | Dyspnoea-12 | Numerical scale | Score >= 12 |
|  | Palpitations | Yes/No | Yes |
|  | Chest pain | Yes/No | Yes |
| Fatigue | FACIT Fatigue scale | Numerical scale | Score < 40 |
| Anxiety/depression* | PHQ-9 | Numerical scale | >= 10 |
|  | GAD-7 | Numerical scale | >8 |
| Cognitive impairment | Montreal Cognitive Assessment (MoCA) | Numerical scale | Score < 26 |
| GI symptoms | Stomach pain | Yes/No | ‘Yes’ to at least 2 symptoms. |
|  | Constipation | Yes/No |  |
|  | Diarrhoea | Yes/No |  |
|  | Weight loss | Yes/No |  |
|  | Nausea and/or vomiting | Yes/No |  |
| Recovered | Do you feel fully recovered from COVID-19? | ‘Yes’ Or below threshold scores to all of above. | |

**Table S1. Clinical questionnaires used to define symptom groups.** The MoCA assessment was administered by a research healthcare worker and the remaining scores were self reported. All non-numerical scores were self-reported and patients were given the option to provide a binary response ‘YES’ or ‘NO’. *Patients were placed in this symptom group if they met the positive threshold for at least one variable in this category.

| Healthy control demographics | | |
| --- | --- | --- |
|  | Nasal fluid (n=25) | BALF (n=9) |
| COVID status | Samples collected prior to COVID-19 pandemic | SARS-COV-2 naïve status confirmed by PCR and serology |
| Age (years) | 39.5 (12.2) | 38.1 (13.3) |
| Female sex | 14 (56) | 2 (22) |
| Non-smoker | 25 (100) | 9 (100) |
| White ethnicity | 14 (56) | 7 (77.8) |

**Table S2.** **Demographics of control participants.** Nasal fluid from convalescent patients were compared to pre-pandemic nasal samples from healthy controls. Sputum from convalescent patients were compared to Broncho-alveolar lavage fluid (BALF) collected from SARS-CoV-2 naive individuals. Data is shown as n (%) or mean (SD).

| **Cardiorespiratory** | **Neuropsychiatric** | **GI** |
| --- | --- | --- |
| Atrial fibrillation | Migraine | Inflammatory Bowel disease |
| Ischaemic heart disease | Previous CVA | Irritable bowel syndrome |
| Congestive heart failure | Dementia | Peptic ulcer disease |
| Coronary heart disease | Multiple sclerosis | Chronic liver disease |
| Valvular heart disease | Depression | Any other chronic gastrointestinal disorder |
| COPD | Anxiety |  |
| Asthma | Previous treatment by mental health professional |  |
| Intersitial lung disease | Any other chronic neurological disorder |  |
| Bronchiectasis |  |  |
| Previous pulmonary emobolus |  |  |
| Previous TB |  |  |
| Lung carcinoma |  |  |
| Any other chronic cardiac or lung disorder |  |  |

**Table S3.** **Comorbidities included as covariates in analysis.** Details of patient questionnaire items used to assign patient comorbidity status relevant to the long COVID symptom outcomes. Relevant comorbidities were included as covariates in the model.

**Supplementary figure legends**

**S1. Univariate analysis of proteins associated each symptom.** A univariate analysis of proteins identified by PLR for each symptom outcome is shown. Wilcoxon signed-rank test was used to compare medians between individuals with long COVID symptoms and those who were recovered. * = p<0·05, ** = p<0·01, *** = p<0·001, ****= p<0·0001.

**S2. Comparison of inflammatory proteins between sexes.** Proteins that were associated with symptoms were compared between men and women who experienced fatigue (A & B), anxiety/depression (C), GI symptoms (D–F) and cognitive impairment (G). Patients were divided into age groups < 50 yrs and >/= 50 years to account for changes in oestogen levels during the menopause which might cause sex differences. Differences between men and women were compared using the Wilcoxon signed-rank test. * = p<0·05, ** = p<0·01, *** = p<0·001, ****= p<0·0001.

**S3. Inflammatory mediators and SARS-CoV-2 antigen in the respiratory tract.** N antigen in sputum from patients who were recovered and those who had long COVID with respiratory symptoms (A). BALF from healthy SARS-COV-2 naïve individuals were used as controls. Medians were compared using the Wilcoxon signed-rank test. The horizontal line denotes the lower limit of detection for the assay. Levels of nasal cytokines were compared between patients with and without cardiorespiratory symptoms (B–G). Nasal cytokines were selected for this analysis if they were found in plasma to be associated with increased odds of cardiorespiratory symptoms. Difference in medians were all non-significant using the Wilcoxon signed rank test. The relationship between plasma and nasal cytokine levels in patients with paired samples collected were compared using Spearman’s rank correlation coefficient (n=70) (H–M).

**S4. PLR classification error and AUC.** A 50 repeats 10-fold nested cross-validation was used to select the predictive model for each symptom outcome, selecting the optimal lambda for each model as well as the model with the lowest classification error. The distribution of classification error (A) and area under curve (AUC) (B) from the nested cross-validation is shown.
